# Optimizing the sensitivity of detection of respiratory syncytial virus infections in longitudinal studies using the combination of weekly sample testing and biannual serology

**DOI:** 10.1101/2025.09.18.25336083

**Authors:** Shannon C. Conrey, Daniel C. Payne, Maria Deza Leon, Monica Epperson, Melissa M. Coughlin, Allison R. Burrell, Claire P. Mattison, Rachel M. Burke, Julia M. Baker, Natalie J. Thornburg, Meredith L. McMorrow, Mary Allen Staat, Ardythe L. Morrow

**Author notes:** **Corresponding author:** Shannon Conrey. Author contributions: Conceptualization: SCC, ALM; data curation: SCC, ARB, CPM; formal analysis: SCC; funding acquisition: ALM, DCP, MAS; investigation: SCC, ALM, DCP, RMB, MDL, ME, MC, JMB, NT, MLM; methodology: SCC, ALM, RMB; project administration: SCC, DCP, MAS, ALM; software: SCC; supervision: ALM, DCP, MAS; writing, original draft: SCC (lead), DCP, ALM; writing-review and editing: SCC, DCP, ME, MMC, CPM, RMB, JMB, NJT, MLM, ALM. All authors reviewed and approved the final manuscript. Funding: The PREVAIL Cohort was funded by a cooperative agreement from the US Centers for Disease Control and Prevention (IP16-004), with additional support provided by the Molecular Epidemiology in Children’s Environmental Health Training program (5 T32 ES 10957-18), the National Center for Advancing Translational Sciences of the National Institutes of Health (UL1TR001425) and the IMPRINT Cohort (U01A1144673). Data analysis and manuscript preparation was funded by the MOM2CHild study (R01HD109915). The findings and conclusions in this report are those of the authors and do not necessarily represent the official position of the CDC or NIH.

## Abstract

Cohort studies are often challenged by incomplete adherence to sampling regimens, limiting the full capture of disease burden. We describe the detection of respiratory syncytial virus (RSV) infections achieved in a birth cohort using a combination of weekly nasal sample testing and serology.

The PREVAIL Cohort followed 245 maternal-child dyads from birth to age 18-24 months. Weekly mid-turbinate nasal swabs were tested for RSV using real-time polymerase chain reaction (RT-qPCR). Serum was tested for RSV pre-fusion F IgG and IgA antibody at age 6 weeks and biannually from 6-24 months. Mixed effects classification and regression trees (CART) identified antibody thresholds consistent with a RT-qPCR-identified RSV infection using a subset of participants having ≥90% weekly sample adherence (*n*=53, 21%). Resulting thresholds were applied to participants with either ≥70% of weekly samples or serum at age 18-24 months (*n*=194, 79%) Incidence rates were compared using Fisher’s exact test.

CART identified a log_10_ change in IgG>0.32 or IgA>0.20 as indicative of an RSV infection. Comparing RT-qPCR-only to a combination of RT-qPCR and serology, RSV cumulative incidence (49% *vs* 75%, *p*<0.001) and incidence density (0.33 *vs* 0.71 infections/child-year, *p*<0.001) increased; these rates did not differ from those calculated in those with ≥90% sample adherence.

**Key messages:**

- We sought to develop a method to maximize RSV infection detection to optimize estimation of disease burden in longitudinal studies, which are prone to incomplete protocol adherence to weekly sample collection.
- Using a combination of weekly sample submissions and regular IgG and IgA serology, we identified incident RSV infections in participants with lower weekly sample submissions.
- The combination of weekly samples and periodic serology can be used to increase power, reduce selection bias, and improve study compatibility in infectious disease cohort studies.

## Introduction

Cohort studies can provide powerful insights into the natural history of acute infections through longitudinal testing and follow-up^1, 2^ . However, complete ascertainment of infections depends on participant adherence to frequent sample collection, which is highly variable^1, 3–5^. Restricting analyses to participants with high sample adherence optimizes infection detection^3, 4, 6^ but reduces sample size and may differentially exclude participants^1^, resulting in potential selection bias^7^. Reducing participant burden by collecting samples only when symptomatic may increase participation^4, 8^ but limits the ability to detect asymptomatic infections, substantially underestimating the burden of infection^9, 10^.

Use of serological assays in cohort studies provides another approach to detecting infections^11^ and may reduce selection and misclassification biases^3^. However, most serologic assays are validated to identify seropositivity^3, 12^, defined as having ever been infected, requiring further evaluation to detect repeat infections and calculate incidence rates^12^. Furthermore, serological detections alone cannot date an infection, and therefore restrict the ability to assess symptomatology, healthcare utilization, or time-varying factors associated with disease^9^.

Recently, two immunization strategies were approved for prevention of RSV in infants^13,14^. Accurately and completely capturing symptomatic and asymptomatic RSV infections before and after immunization introduction is essential to assessing vaccine effectiveness and immune response^10^. However, the lack of consistent methods to determine RSV incidence results in heterogeneity and lack of comparability across studies^9, 15–18^. Here, we describe an integrated method to identify incident RSV infections using a combination of real-time polymerase chain reaction (RT-qPCR)-tested nasal swabs and regular serologic collection in a two-year birth cohort. We compare detection sensitivity by methodologic decision points and evaluate the impact of methods on statistical power, selection and misclassification bias.

## Methods

### Study cohort

The Pediatric Respiratory and Enteric Viral Acquisition and Immunogenesis Longitudinal (PREVAIL) Cohort is a two-year birth cohort conducted in Greater Cincinnati, Ohio from April 2017-July 2020. Institutional review board approvals were obtained from the US Centers for Disease Control and Prevention, Cincinnati Children’s Hospital Medical Center, and enrolling birth hospitals. Full study methods have been published^2^. Briefly, prenatal enrollment included mothers who were ≥34 weeks of gestation, ≥18 years of age, and expecting a healthy, singleton child. Children were followed from birth until age two, including weekly symptom surveillance *via* text message, weekly nasal swab and stool sample submission, and regular in-person study visits and blood draws.

### RSV detection

Mothers collected mid-turbinate nasal samples each week using flocked swabs (Copan Diagnostics, Inc.), placed into vials containing BD Universal Viral Transport medium (Becton, Dickinson, and Co., Franklin Lakes, NJ) and delivered to the study laboratory within 48 hours of collection. Swabs were tested for respiratory pathogens, including RSV A and B, using the Luminex multiplex RT-qPCR NxTAG Respiratory Pathogen Panel assay^19^. Detections were considered part of a continuous RSV infection if the same subtype was detected ≤30 days apart^9^. RT-qPCR-detected infections were dated using the date of the first RT-qPCR-positive.

Serum samples were collected using a standardized protocol at birth and at in-clinic study visits at ages 6 weeks and 6, 12, 18, and 24 months. Serum antibodies (IgA and IgG) responsive to RSV were measured using the MesoScale Discovery Respiratory Panel 1 multiplex serology assay (MesoScale Diagnostics, Rockville MD) according to the manufacturer’s instructions, calculated in arbitrary units (AU) per mL using standard curves with manufacturer provided calibrators. Concentrations below or above the level of quantification (LOQ) were imputed as half the lower LOQ or the upper LOQ, respectively.

### Definition of adherence levels

To illustrate the differences in power and bias associated with restricting inclusion criteria by adherence level, we identified and compared subsets using varying weekly sample adherence levels as inclusion criteria (Figure 1). It is important to note that each of these rules is applied to the prior subset, so the subsets are nested. Participants were considered evaluable for infectious outcomes using a combination of RT-qPCR and serology if they participated in the study ≥18 months and submitted either ≥70% of weekly samples or provided a serum sample at ≥18 months of age^2^. For incidence calculations using RT-qPCR only, we restricted analysis to evaluable participants who were ≥70% weekly sample adherent^9, 20, 21^. For analysis requiring minimal gaps in submissions, we restricted analysis to participants who submitted ≥90% of weekly samples.

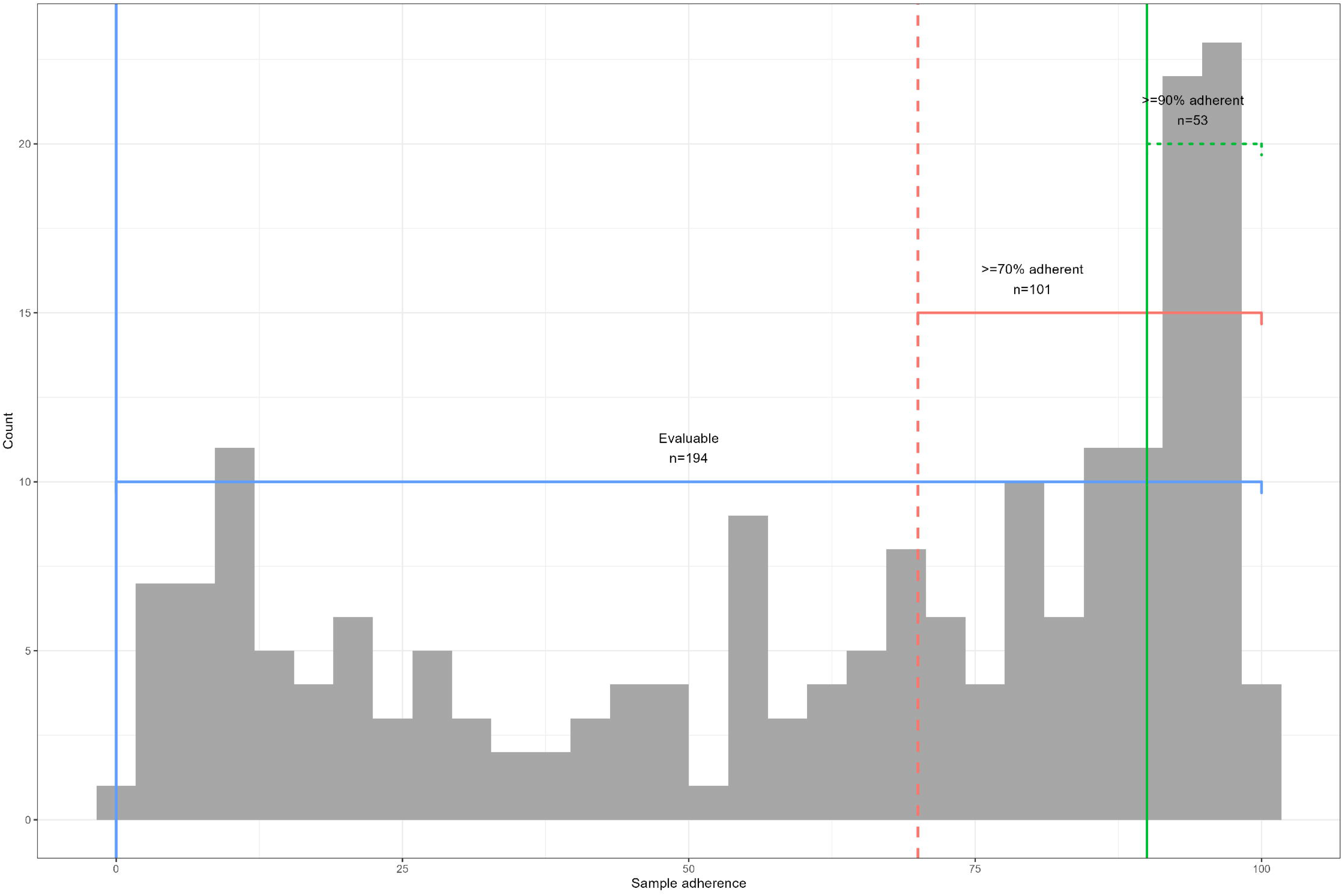

### Identification of thresholds

Using the ≥90% adherent subset, we applied a mixed-effects Classification and Regression Tree (CART) analysis to generate detection thresholds in serum antibody consistent with having an incident RSV infection during the interim between blood draws (study interval). CART models consider all possible combinations and levels of variables to recursively split data, selecting thresholds in the available predictors that result in the most accurate terminal nodes^22^.

Study intervals were identified as positive or not positive for RSV based on the presence of a RT-qPCR-identified RSV infection within the interval. Based on the time required to mount a detectable antibody response after an infection, RT-qPCR detections within one month of the blood draw were identified as positive for the subsequent blood draw. Using the change in antibody levels between the start and the end of a study interval, three possible predictors were included in the CART models; 1.) change in log_10_ concentration, 2.) fold-change in concentration, and 3.) a binary indicator of ≥4-fold change in concentration. A fourth predictor, log_10_ concentration at the time of the blood draw, was also included.

Using the ≥90% adherent subset, CART models were constructed for each isotype to produce thresholds for each variable individually, with the interval outcome as positive or not positive and participant ID as the clustering variable. These thresholds were applied and percent agreement was calculated and compared to the validated laboratory-determined receiver operating characteristic (ROC) curves^23^ for seropositivity and to each other for detecting incident infections.

To identify the predictors and thresholds that resulted in the most accurate identification of incident infection, a combined CART model for each isotype was constructed using all four variables to identify without restriction to the number of splits or application of variable weights. Using the resulting thresholds, serology-identified positives were assigned the date of the serum sample in which the positive was detected. Serologic positives associated with a RT-qPCR-positive study interval were considered confirmatory of the RT-qPCR-positive, while serologic positives without a RT-qPCR-positive in the study interval were considered new incident infections. Investigations, including timing of the serum sample, weekly sample adherence, and reported respiratory symptoms during the study interval, were conducted for all unconfirmed RT-qPCR-identified infections and new incident infections. Once detection thresholds were established in ≥90% adherent participants, they were applied to all evaluable participants.

### Statistical analysis

Fisher’s exact test or Kruskal-Wallis were used to compare socio-demographic factors among the adherence-defined subsets to illustrate selection bias introduced with increasing adherence level. Cumulative incidence in each subset was calculated for infections detected using RT-qPCR only and by combining RT-qPCR and serology. To calculate incidence density, weeks-at-risk was calculated by multiplying the number of infections by four weeks, as RSV detections within 30 days were considered part of a continuous infection^9^, then subtracting the resulting number from the total weeks of follow-up. Fisher’s Exact test with Holms corrections for multiple comparisons was used to compare proportions of infections identified by RT-qPCR and by serology, as well as cumulative incidence and incidence density for each subset.

To evaluate the effect of study design criteria on power, we calculated detectable effect size at 80% power for a chi-squared test for each of our three adherence subsets. To examine the impact on misclassification and selection biases, we compared the demographic factors between those RSV-infected *vs* not RSV-infected in our evaluable subset using the RT-qPCR-only method, serology-only method, and the combined method using Kruskal-Wallis for continuous and Fisher’s Exact Test for categorical data. All analyses were conducted using the R Environment for Statistical Computing (version 4.2.3)^22, 24^.

## Results

PREVAIL enrolled 245 participants; 194 (79%) were defined as evaluable, 101 (41%) were ≥70% adherent, and 53 (22%) were ≥90% adherent (Figure 2). Each increase in adherence resulted in a subset with greater proportions of subjects who identified as White race, higher income, married, privately insured, higher education level and who breastfed for longer durations (Table 1).

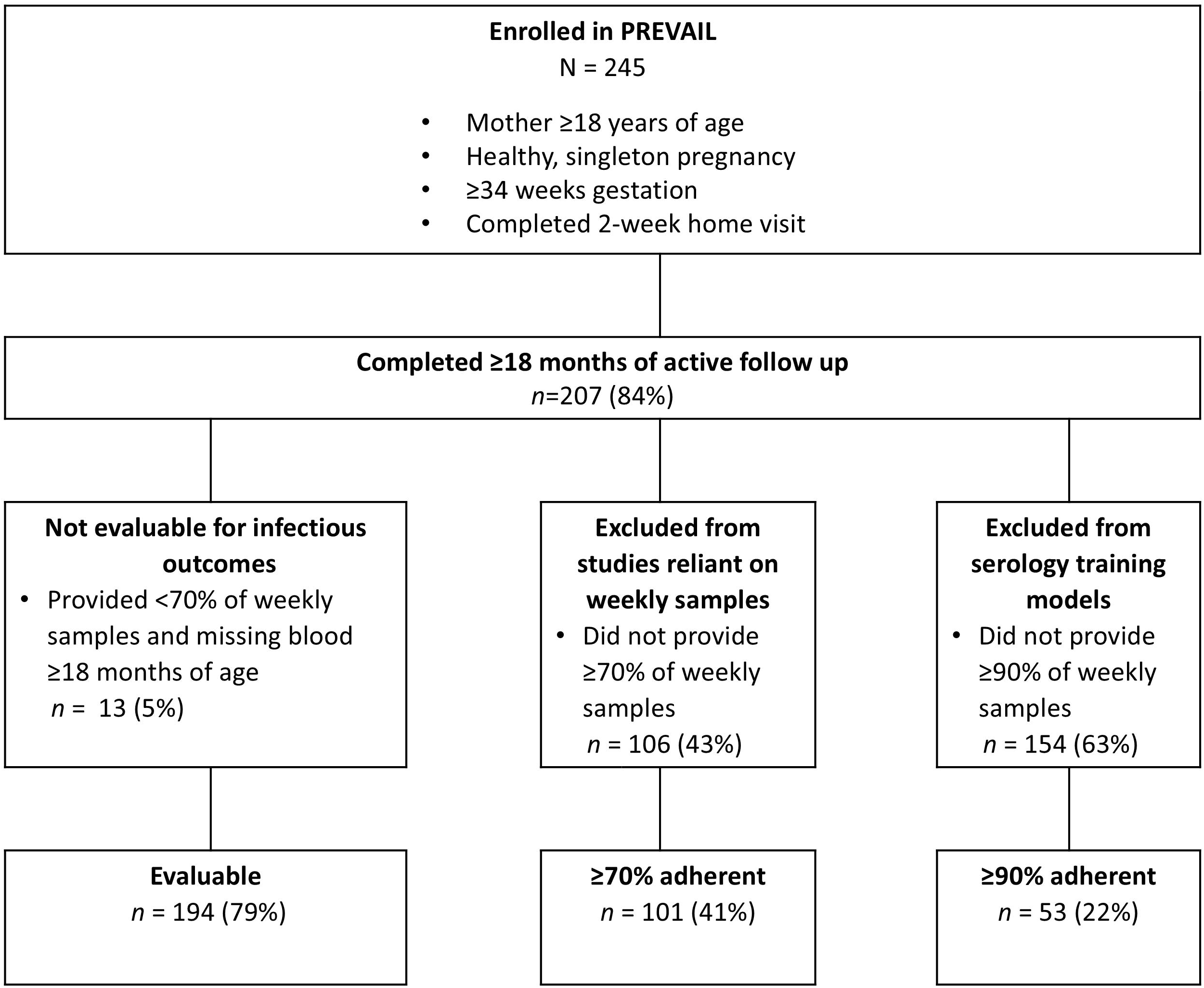

**Table 1:** demographics by subset inclusion criteria.

|  |  | Enrolled in<br>PREVAIL<br>N = 245<sup>1</sup> | Evaluable<sup>a</sup><br>n = 194<sup>1</sup> | p<sup>2</sup> | ≥70%<br>adherent<sup>*</sup><br>n = 101<sup>1</sup> | p<sup>2</sup> | ≥90%<br>adherent<sup>§</sup><br>n = 53<sup>1</sup> | p<sup>2</sup> |
| --- | --- | --- | --- | --- | --- | --- | --- | --- |
| **Nasal swab adherence** |  | 61% (22, 88) | 71% (30, 91) | <0.001 | 91% (81, 94) | <0.001 | 94% (91, 95) | <0.001 |
| **Maternal age** | Years | 29.6 (25.7, 33.2) | 30.2 (26.9, 33.7) | <0.001 | 32.6 (29.1, 34.9) | <0.001 | 33.0 (30.0, 34.6) | <0.001 |
| **Race** | Black | 107 (44%) | 82 (42%) | 0.5 | 17 (17%) | <0.001 | 6 (11%) | <0.001 |
|  | White/Other | 138 (56%) | 112 (58%) |  | 84 (83%) |  | 47 (89%) |  |
| **Hispanic Ethnicity** | Yes | 6 (2%) | 6 (3%) | 0.3 | 5 (5%) | 0.084 | 3 (6%) | 0.20 |
| **Insurance type** | Private | 106 (43%) | 93 (48%) | 0.004 | 78 (77%) | <0.001 | 48 (91%) | <0.001 |
|  | Public | 139 (57%) | 101 (52%) |  | 23 (23%) |  | 5 (9.4%) |  |
| **Marital Status** | Married/Parter | 163 (67%) | 134 (69%) | 0.10 | 87 (86%) | <0.001 | 47 (89%) | <0.001 |
|  | Single | 82 (33%) | 60 (31%) |  | 14 (14%) |  | 6 (11%) |  |
| **Maternal Education** | ≤HS | 115 (47%) | 81 (42%) | 0.002 | 18 (18%) | <0.001 | 5 (9.4%) | <0.001 |
|  | >HS | 130 (53%) | 113 (58%) |  | 83 (82%) |  | 48 (91%) |  |
| **Family Income** | <\$25k | 88 (36%) | 61 (31%) | 0.001 | 12 (12%) | <0.001 | 2 (3.8%) | <0.001 |
| | \$25k-\$50k | 49 (20%) | 36 (19%) | | 10 (10%) | | 3 (5.7%) | |
| | >\$50k | 108 (44%) | 97 (50%) | | 79 (78%) | | 48 (91%) | |
| **Infant sex** | Female | 127 (52%) | 96 (49%) | 0.2 | 46 (46%) | 0.10 | 22 (42%) | 0.14 |
|  | Male | 118 (48%) | 98 (51%) |  | 55 (54%) |  | 31 (58%) |  |
| **Breastfeeding initiation** |  | 212 (87%) | 171 (88%) | 0.15 | 96 (95%) | 0.001 | 51 (96%) | 0.03 |
| **Duration of BF** | days | 83 (16, 312) | 109 (20, 379) | 0.001 | 297 (66, 429) | <0.001 | 335 (162, 428) | <0.001 |
| **Delivery mode** | C-Section | 94 (38%) | 78 (40%) | 0.2 | 48 (48%) | 0.014 | 25 (47%) | 0.30 |
|  | Vaginal | 151 (62%) | 116 (60%) |  | 53 (52%) |  | 28 (53%) |  |
| **Birthweight (kg)** |  | 3.29 (2.95, 3.53) | 3.29 (2.98, 3.55) | 0.3 | 3.33 (3.09, 3.61) | 0.011 | 3.37 (3.09, 3.61) | 0.20 |
| <sup>1</sup> median (IQR) or n (%) |  |  |  |  |  |  |  |  |
| <sup>2</sup> Kruskal Wallis or Fisher exact test, based on difference by subset inclusion criteria compared to enrolled cohort |  |  |  |  |  |  |  |  |
| Adherence categories were all subsets of the 245 enrolled participants; thus each subset was nested within the larger groupings. The p-values represent the difference between those who meet each adherence category and all enrolled participants. |  |  |  |  |  |  |  |  |
| <sup>a</sup>Evaluable participants were under active follow-up for at least 18 months and either submitted ≥70% of weekly samples or provided a serum sample ≥18 months of age |  |  |  |  |  |  |  |  |
| <sup>*</sup>≥70% adherent participants were under active follow-up for at least 18 months and submitted ≥70% of weekly samples. To minimize missed infections, this group is used for analysis using only RT-qPCR-identified infections |  |  |  |  |  |  |  |  |
| <sup>§</sup>≥90% adherent participants were under active follow-up for at least 18 months and submitted ≥90% of weekly samples. This group is used when true positives and negatives are required |  |  |  |  |  |  |  |  |

### Identification of RSV thresholds in the ≥90% adherent participants

RT-qPCR identified 54 RSV infections in 36 children (68%) in the ≥90% adherent subset, with an incidence density of 0.55 infections/child-year. While the CART-identified thresholds (Table 2) for the three predictors related to change in antibody had high percent agreement in identifying incident infections, the threshold selected for log_10_ concentration resulted in a high number of serial positives, overestimating incident infections. Comparisons between the validated ROC curves and the CART-derived thresholds were completed using only IgA, as all children were seropositive using the ROC curves for IgG between birth and month 6. Each of the four thresholds had high percent agreement for identifying seropositivity compared to the ROC analysis., ranging from 88.5%-96.2%. In the combined model, CART identified a single variable and threshold per isotype, log_10_ increases of >0.202 or >0.32 AU, for IgA and IgG, respectively, from the previous blood draw as the most accurate predictor of an incident RSV infection, hereafter referred to concentration change.

**Table 2:**
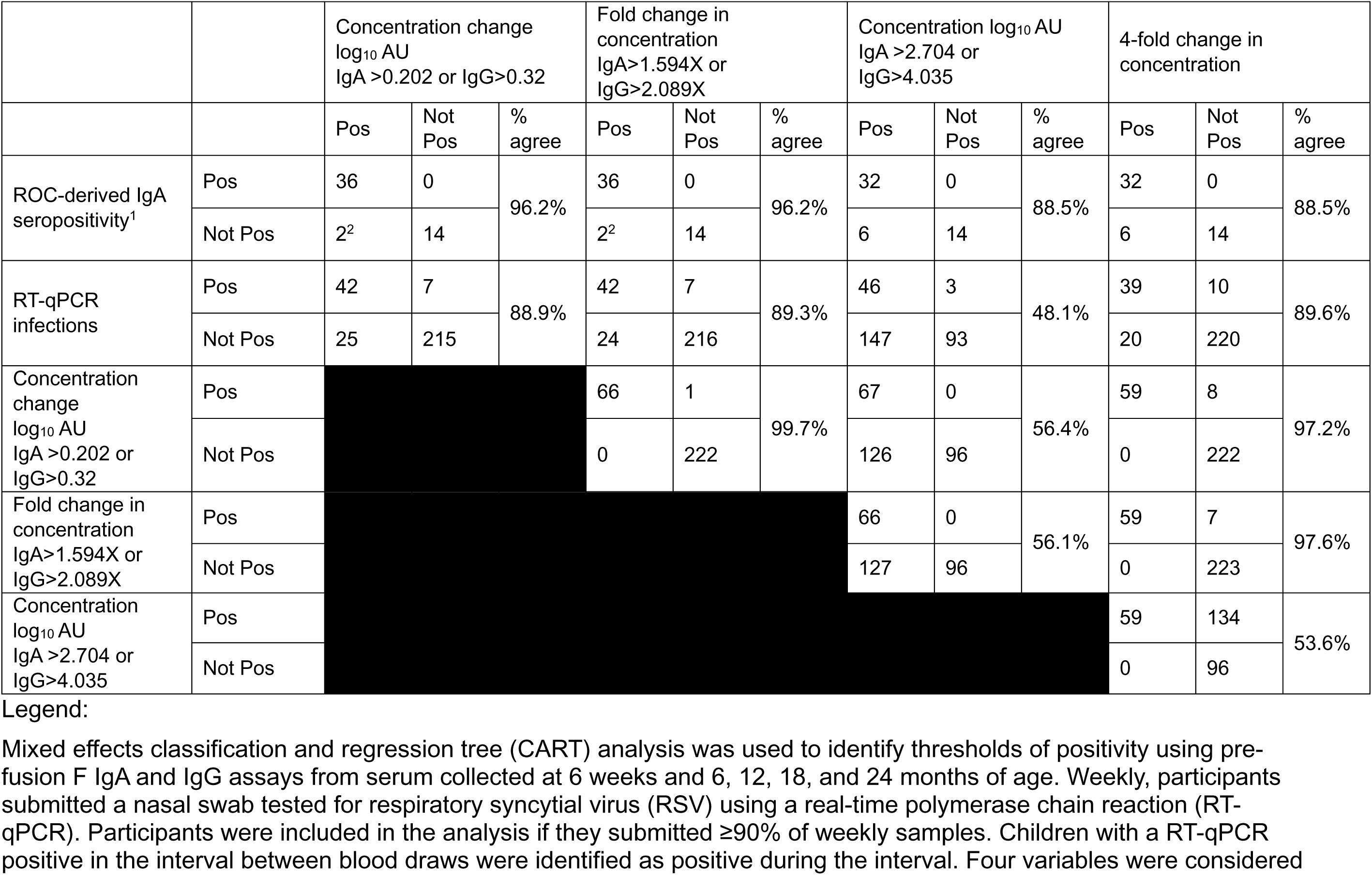
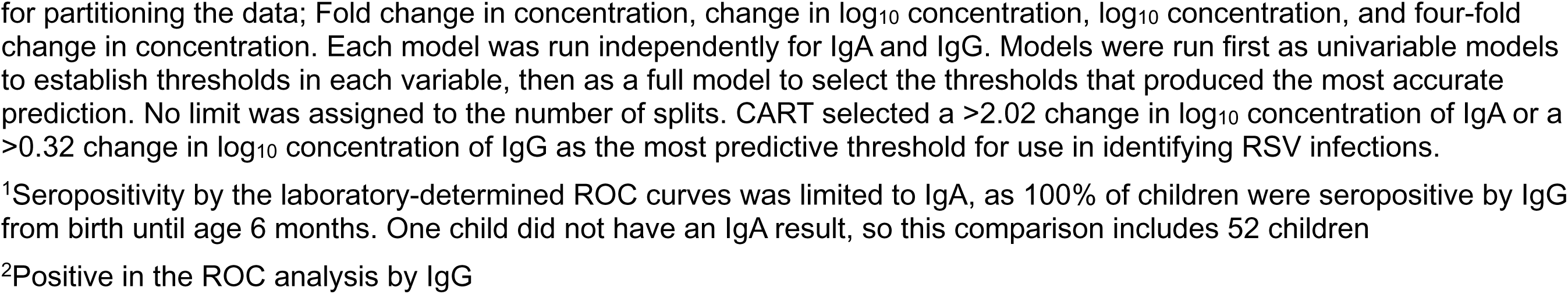
Comparison between methods to identify RSV infections in 53 highly-adherent PREVAIL children.

|  |  | Concentration change<br>log<sub>10</sub> AU<br>IgA >0.202 or IgG>0.32 |  |  | Fold change in<br>concentration<br>IgA>1.594X or<br>IgG>2.089X |  |  | Concentration log<sub>10</sub> AU<br>IgA >2.704 or<br>IgG>4.035 |  |  | 4-fold change in<br>concentration |  |  |
| --- | --- | --- | --- | --- | --- | --- | --- | --- | --- | --- | --- | --- | --- |
|  |  | Pos | Not<br>Pos | %<br>agree | Pos | Not<br>Pos | %<br>agree | Pos | Not<br>Pos | %<br>agree | Pos | Not<br>Pos | %<br>agree |
| ROC-derived IgA<br>seropositivity<sup>1</sup> | Pos | 36 | 0 | 96.2% | 36 | 0 | 96.2% | 32 | 0 | 88.5% | 32 | 0 | 88.5% |
|  | Not Pos | 2<sup>2</sup> | 14 |  | 2<sup>2</sup> | 14 |  | 6 | 14 |  | 6 | 14 |  |
| RT-qPCR<br>infections | Pos | 42 | 7 | 88.9% | 42 | 7 | 89.3% | 46 | 3 | 48.1% | 39 | 10 | 89.6% |
|  | Not Pos | 25 | 215 |  | 24 | 216 |  | 147 | 93 |  | 20 | 220 |  |
| Concentration<br>change<br>log<sub>10</sub> AU<br>IgA >0.202 or<br>IgG>0.32 | Pos |  |  |  | 66 | 1 | 99.7% | 67 | 0 | 56.4% | 59 | 8 | 97.2% |
|  | Not Pos |  |  |  | 0 | 222 |  | 126 | 96 |  | 0 | 222 |  |
| Fold change in<br>concentration<br>IgA>1.594X or<br>IgG>2.089X | Pos |  |  |  |  |  |  | 66 | 0 | 56.1% | 59 | 7 | 97.6% |
|  | Not Pos |  |  |  |  |  |  | 127 | 96 |  | 0 | 223 |  |
| Concentration<br>log<sub>10</sub> AU<br>IgA >2.704 or<br>IgG>4.035 | Pos |  |  |  |  |  |  |  |  |  | 59 | 134 | 53.6% |
|  | Not Pos |  |  |  |  |  |  |  |  |  | 0 | 96 |  |

Mixed effects classification and regression tree (CART) analysis was used to identify thresholds of positivity using pre-fusion F IgA and IgG assays from serum collected at 6 weeks and 6, 12, 18, and 24 months of age. Weekly, participants submitted a nasal swab tested for respiratory syncytial virus (RSV) using a real-time polymerase chain reaction (RT-qPCR). Participants were included in the analysis if they submitted ≥90% of weekly samples. Children with a RT-qPCR positive in the interval between blood draws were identified as positive during the interval. Four variables were considered
for partitioning the data; Fold change in concentration, change in $\log_{10}$ concentration, $\log_{10}$ concentration, and four-fold change in concentration. Each model was run independently for IgA and IgG. Models were run first as univariable models to establish thresholds in each variable, then as a full model to select the thresholds that produced the most accurate prediction. No limit was assigned to the number of splits. CART selected a $>2.02$ change in $\log_{10}$ concentration of IgA or a $>0.32$ change in $\log_{10}$ concentration of IgG as the most predictive threshold for use in identifying RSV infections.
<sup>1</sup>Seropositivity by the laboratory-determined ROC curves was limited to IgA, as 100% of children were seropositive by IgG from birth until age 6 months. One child did not have an IgA result, so this comparison includes 52 children
<sup>2</sup>Positive in the ROC analysis by IgG

After applying the concentration change thresholds to our training data (Figure 3A), we identified 21 previously undetected RSV infections in 17 children, including 4 infections in 3 children with no detectable RT-qPCR-identified infection (Figure 3B). Most (*n*=19, 90%) of these new infections were identified using IgA (Supplemental Table 2); 33% (*n*=7) were identified using IgA only, including both new infections in the first 6 months of life. There were 16 repeat infections identified by concentration change; 14 (88%) of these were identified using IgA, with 31% (*n*=5) identified using IgA only. Nearly half of new infections (*n*=10, 48%) were detected following a study interval with very high weekly sample adherence (mean 98.0% ±2.8), while 52% (*n*=11) were detected following study intervals of lower adherence (range 50-88%, mean 64.7%, ±14.6).

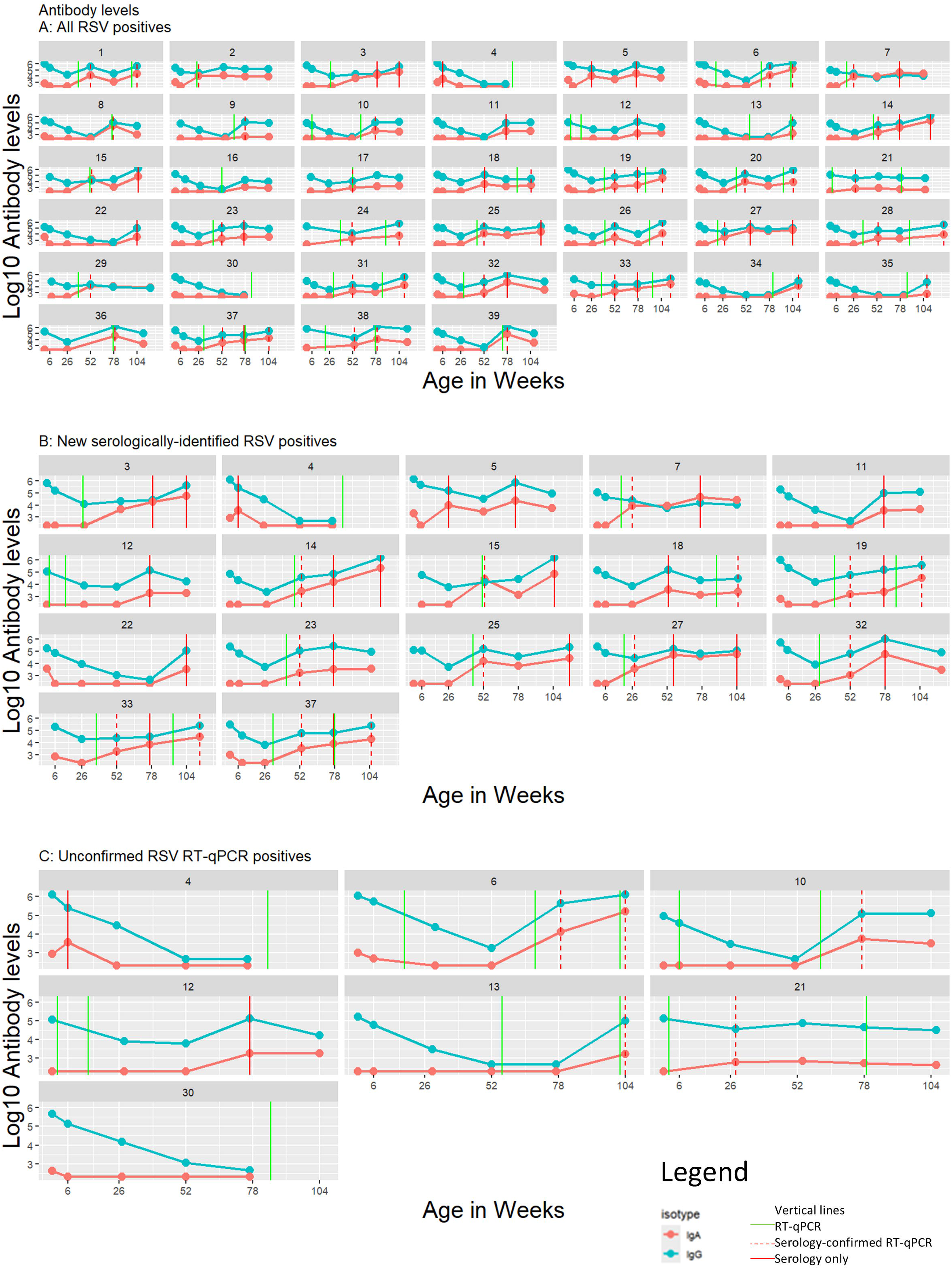

Nearly all children (15/17, 88%) reported respiratory symptoms by text survey during the study interval prior to the serologic positive.

Of the 56 RT-qPCR-identified infections, 48 (86%) were confirmed by serology (Figure 2C). Of the eight unconfirmed, two children did not have a blood draw after the RT-qPCR-positive and five, occurring from 6 to 79 weeks of age, were not associated with any symptoms. The final infection, occurring at two weeks of age, was symptomatic and resulted in an emergency department visit. Although none of these children were seropositive using either ROC or concentration change, given the high specificity of the Luminex NxTAG RPP (1, 95%CI 0.97-1)^25^, these infections were retained for incidence calculations. After combining RT-qPCR-and concentration change-identified infections, we identified 77 infections in 39 children, resulting in 74% cumulative incidence and 0.77 infections/child-year (Figure 4).

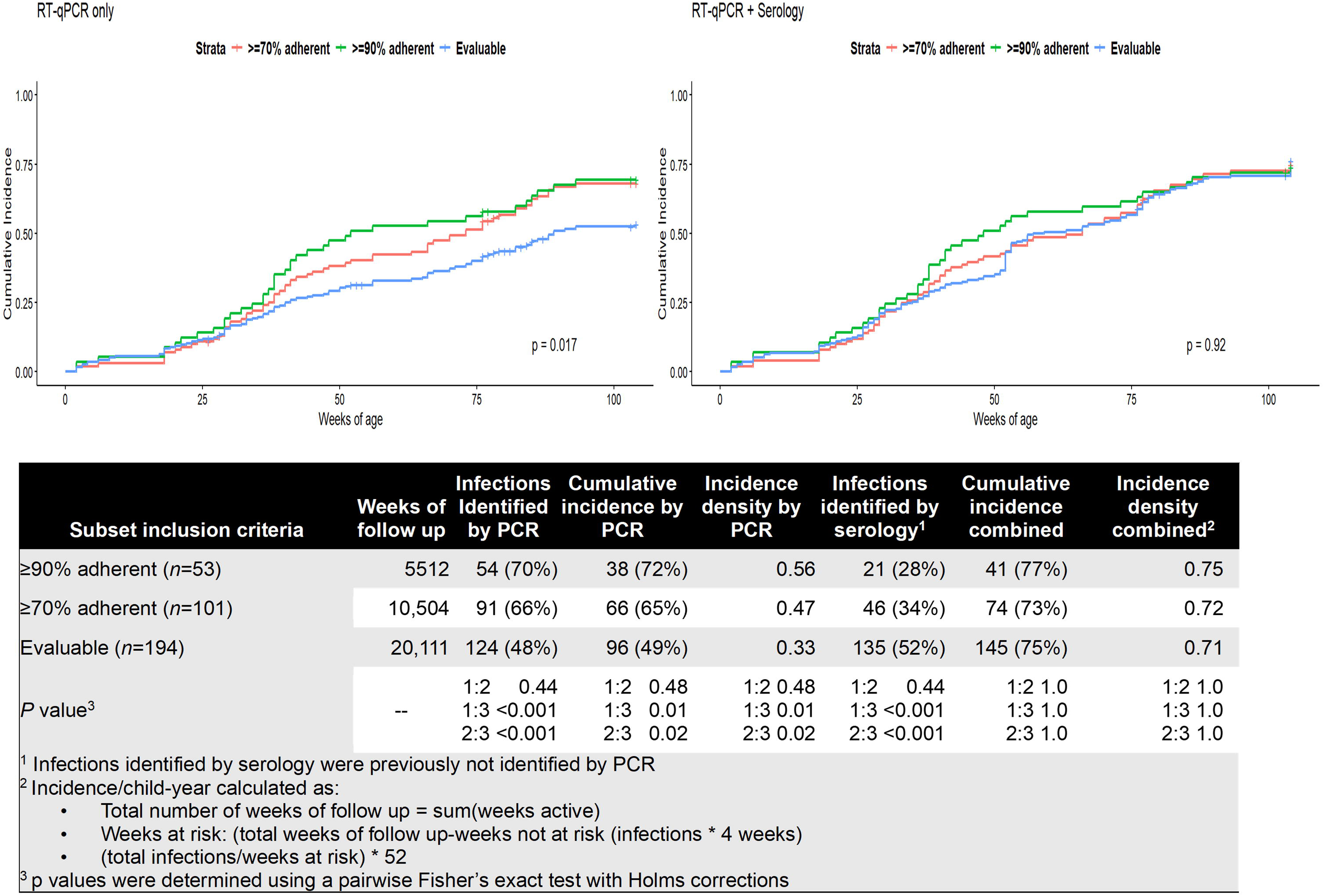

### Application to all evaluable participants

RT-qPCR detected 124 RSV infections in 96/194 (49%) children, with an incidence density of 0.33 infections/child-year (Figure 4). Concentration change identified 133 new infections, including 85 infections in 49 children who previously did not have a RT-qPCR-detected infection (Supplemental Table 2); 51/133 (38%) were identified using IgA only, 3/133 (2%) were identified using IgG only, and 79/133 (60%) met both criteria. All new infections in the first year of life (*n*=11) were identified using IgA only. Nearly all repeat infections (73/76, 96%) were identified using IgA, 39% (30/76) using IgA only. All three IgG-only infections were repeat infections in the second year of life.

Of the 124 RT-qPCR-identified infections, 109 (88%) were confirmed serologically; nearly half (7/15, 47%) of the unconfirmed RT-qPCR infections occurred in the first 6 months of life and 53% (8/15) were asymptomatic. When combining RT-qPCR and serology, 259 infections were identified in 145 children, increasing cumulative incidence to 75% by two years of age (*p*<0.001), and incidence density to 0.71 infections/child-year (*p*<0.001).

### Comparison of incidence, power achieved, and bias by protocol adherence and method

Using RT-qPCR-identified infections, cumulative incidence at 2 years of age (72%, 65%, 49%, *p*=0.02) and infections/child-year (0.56, 0.47, 0.33, *p*=0.003) varied among the ≥90% adherent, ≥70% adherent, and the Evaluable group, respectively. (Figure 4). Using our integrated approach, no differences remained in cumulative incidence at 2 years of age (77%, 73%, 75%, *p*=0.68) or incidence/child-year (0.77, 0.72, 0.71, *p*=0.20)

In comparing power achieved using chi-squared tests by subset (Figure 5), ≥80% power was achieved with an effect size ≥39% for the ≥90% adherent, ≥28% for the ≥70% adherent, and >20% for all eligible participants.

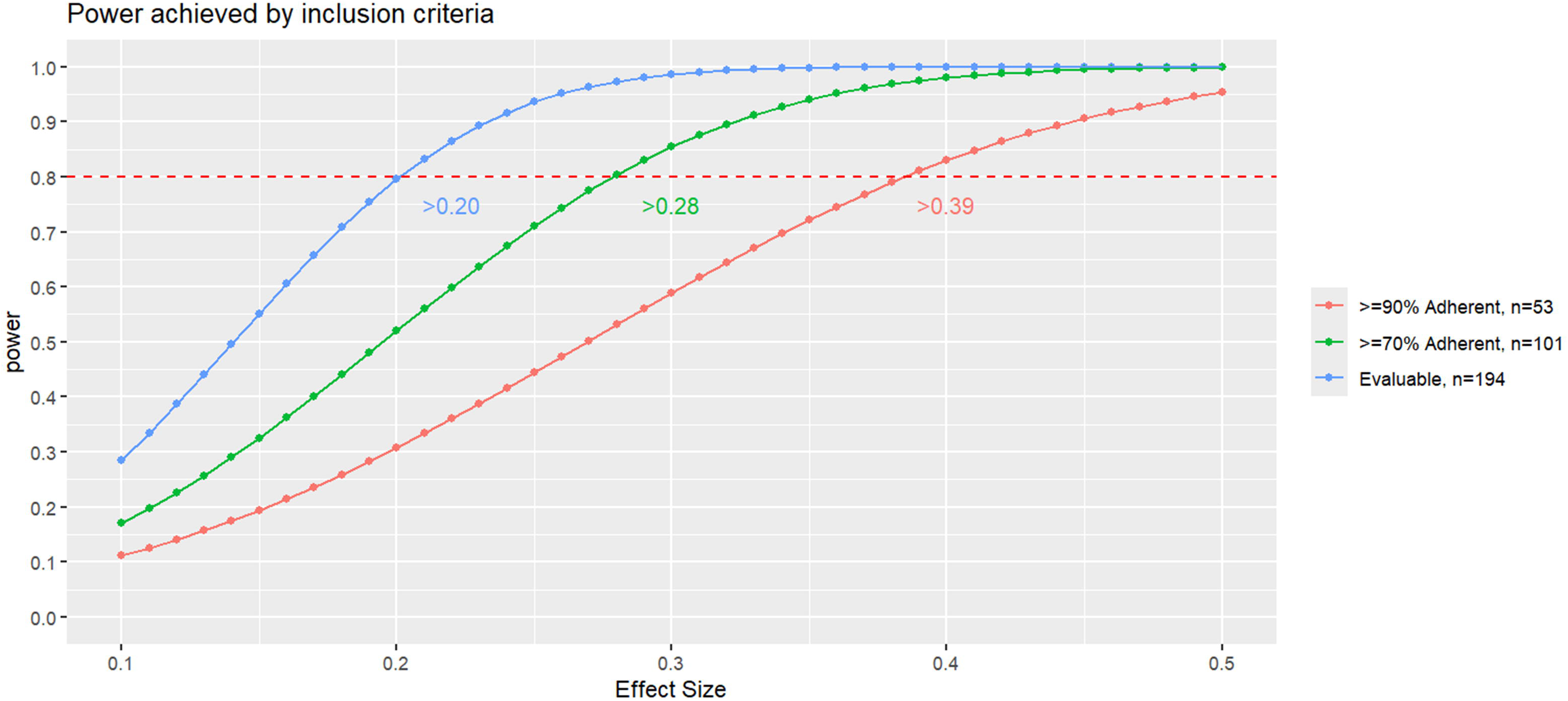

When we compared those that did and did not have a RT-qPCR-identified RSV infection (Table 3), differences by race, insurance type, marital status, maternal education, and family income were detected. However, using all RT-qPCR- and serology-identified infections and comparing those with and without an RSV infection, no differences remained in these factors.

**Table 3:**
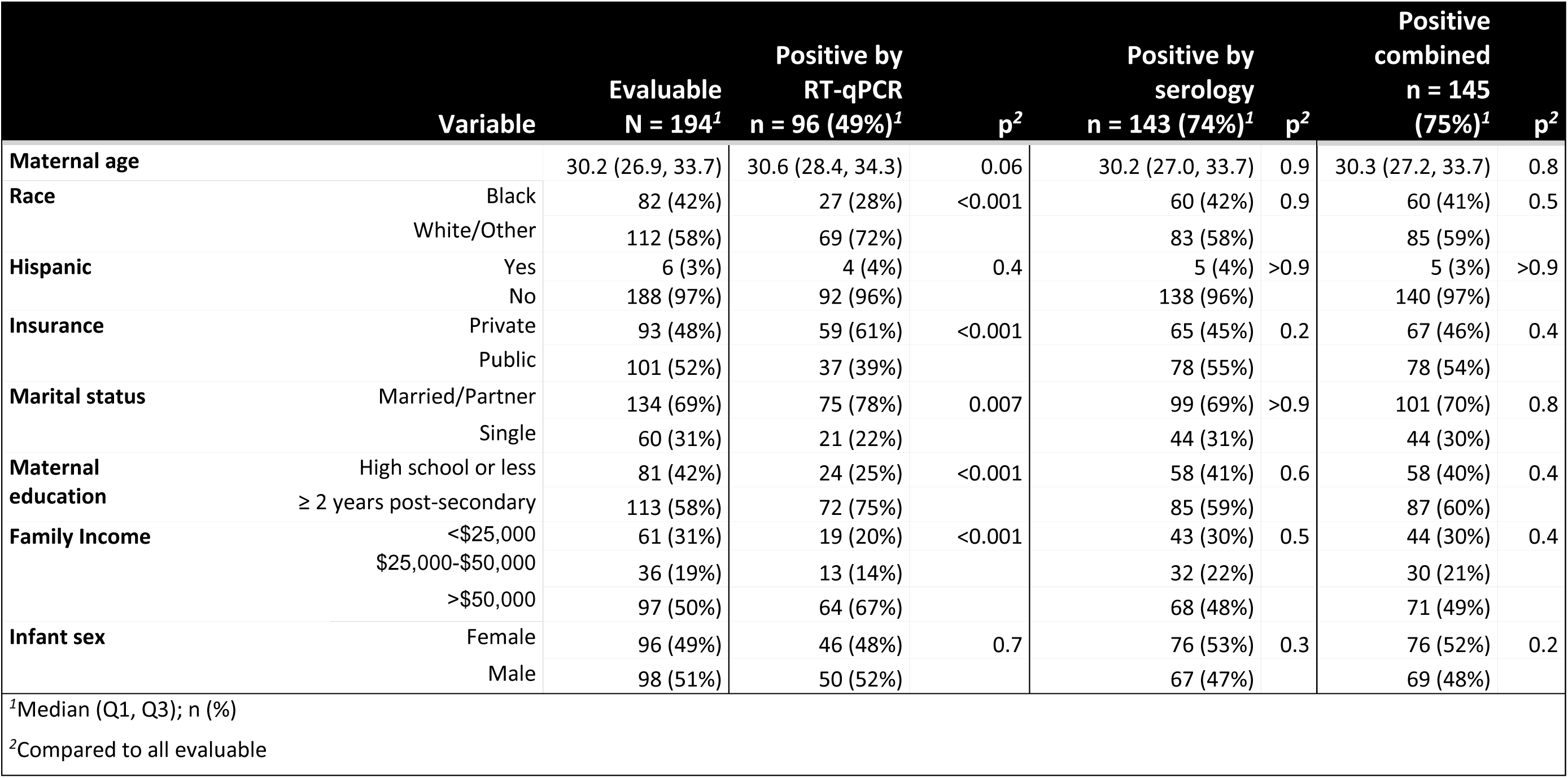
Comparison of RSV infections by demographics using RT-qPCR-only and a combination of RT-qPCR- and serology-identified infections among all evaluable participants.

Using RT-qPCR-only, 49/194 (25%) of evaluable children were misclassified as never having an RSV infection by age two. Furthermore, 25 RT-qPCR-identified infections were misclassified as first infections; 19 of these were reclassified as second and 6 were reclassified as third after adding concentration change-identified infections.

### Sensitivity analysis

Unconfirmed RT-qPCR-positives were removed from incidence rate calculations for each adherence subset (≥90% adherent, ≥70% adherent, evaluable). The adjusted incidence rates (0.71, 0.65, 0.66, respectively) did not differ from each-other (*p*=0.20) or from the previous estimates (*p*=0.22).

## Discussion

The PREVAIL Cohort accurately and more completely identified RSV infections in the first two years of life by integrating RT-qPCR-tested nasal swabs with regular blood draws assayed for both anti-RSV IgG and IgA antibodies. We demonstrate that this integrated method improved our RSV incidence estimates while simultaneously increasing power and reducing selection and misclassification biases. In addition, this method allows comparisons across multiple study designs, including those that rely on RT-qPCR-only and serology-only procedures to detect RSV infections.

Our findings ranging from 0.33-0.56 RSV infections/child-year detected using only RT-qPCR, and 73%-77% cumulative incidence using our integrated approach (Figure 4) are consistent with findings from other international cohorts. The Australian ORChID Cohort reported an incidence rate of 0.46 infections/child-year using weekly RT-qPCR detections only^18^, a retrospective Kenyan cohort reported RSV seropositivity of 83% by age two in all children admitted to a rural Kenyan hospital from 2007-2010^17^, while a prospective Finnish cohort reported 68% seropositivity by age two in healthy children^16^. While the heterogeneity of methods across these studies makes direct comparisons difficult, our findings are consistent with these studies, suggesting that RSV incidence may be more homogeneous world-wide than previously reported^26^ and highlighting the value of a multi-pronged approach to detections^11^.

Four-fold change in antibody has long been used in serology studies to identify new RSV infections^27, 28^. While the percent agreement between our concentration change method and four-fold change was 97.2%, the concentration change method identified five more incident infections missed by RT-qPCR in our highly adherent subset and had higher percent agreement with the ROC-derived seropositivity curves (Table 2).

The durability of IgG combined with its transplacental transfer leads to challenges in identifying early and repeat RSV infections when only IgG is measured^9, 16, 17^. We previously applied CART analysis using the PREVAIL ≥90% adherent subset to identify thresholds in IgG indicative of infection with the endemic coronaviruses OC43, NL63 and HKU1^20^. While the resulting thresholds had high specificity (91.8%, 95.9%, and 90.2%, respectively), transferred maternal IgG antibodies prevented the identification of infections prior to 6 months of age and the durability of IgG prevented the serologic detection of repeat infections^17, 20^. In this analysis, all early and most repeat infections were detected using IgA, many using IgA only, highlighting the need for both IgA and IgG assays for accurate assessment.

After combining RT-qPCR-and serology-detected infections, differences in cumulative incidence and incidence rates by adherence-defined subsets were attenuated, allowing for the inclusion of nearly 80% of enrolled participants and increasing power. In addition, the reduction in selection bias associated with socio-demographic factors using out integrated approach may provide a more robust estimation of effectiveness of prophylactic and therapeutic measures^1, 3, 5, 6^. Finally, the reduction in misclassifications is instrumental for understanding the natural history of immune response^2^.

Although the estimates using concentration change only were nearly identical to those combining both methods, serologically-identified infections cannot be precisely dated, hindering examinations of severity or symptomatology of infections. Such examinations are important for RSV infections, as reinfections are common throughout life^16, 17, 26, 29^. Gauging effectiveness of immunization strategies to prevent disease or reduce severity of symptoms requires precise dating of the immunization event, infection, and symptom onset.

Our study is not without limitations. While the children in the ≥90% adherent group had very high weekly sample submission rates and rarely missed submissions in consecutive weeks, a weekly sample protocol can miss short, transient infections.

Furthermore, weekly sample adherence was calculated over the study duration, but some participants experienced periods of lower adherence, including half of those with serologically-identified new infections. As these participants provided the training data for the CART models, missed infections may have resulted in a misclassification of study intervals as ‘not positive’ and thereby reduced the sensitivity of the detections. In addition, our thresholds are based on a very small subset of our study and should not be considered generalizable to all children.

Our study also has several important strengths. Using CART to identify thresholds using highly adherent children rather than relying on a standardized 4-fold rise increased our detections, while the availability of both IgA and IgG concentrations allowed for the detection of early and repeat infections. While our sample size was modest, our method allowed us to maximize power by including participants with lower weekly sample adherence. Finally, our method reduced potential socio-demographic biases associated with limiting analysis to highly-adherent participants, resulting in greater study robustness and power.

## Conclusions

Using CART analysis to identify serologic thresholds consistent with RSV infection integrated with weekly RT-qPCR-detections allowed the full capture of RSV infections. This approach facilitated improved accuracy of disease burden estimates, a better understanding of RSV immunologic events in infancy and early childhood, and precise dating of infections to determine symptoms status. While larger studies are needed to establish generalizable serology thresholds for RSV infection, use of IgG and IgA serology combined with weekly nasal swab testing is a way for cohort studies to maximize power and reduce biases in RSV detections.

## Data Availability

All data produced in the present study are available upon reasonable request to the authors

---

**Supplemental table 1:** comparison of demographics by subset inclusion criteria.

|  | Overall<br>N = 245<sup>1</sup> | Excluded from<br>analysis<br>N = 51<sup>1</sup> | <70% weekly<br>samples,<br>but serum<br>≥18M<br>N = 93<sup>1</sup> | 70% ≤ weekly<br>samples <90%<br>N = 48<sup>1</sup> | ≥90%<br>weekly samples<br>N = 53<sup>1</sup> | p-value<sup>2</sup> |
| --- | --- | --- | --- | --- | --- | --- |
| **Nasal swab adherence** | 61 (22, 88) | 29 (13, 54) | 29 (11, 51) | 80 (74, 87) | 94 (91, 95) | <0.001 |
| **Maternal age** | 29.6 (25.7, 33.2) | 26.1 (23.2, 30.3) | 28.7 (25.2, 31.3) | 31.6 (28.3, 35.0) | 33.0 (30.0, 34.6) | <0.001 |
| **Race** |  |  |  |  |  | <0.001 |
| Black | 107 (44%) | 25 (49%) | 65 (70%) | 11 (23%) | 6 (11%) |  |
| White/Other | 138 (56%) | 26 (51%) | 28 (30%) | 37 (77%) | 47 (89%) |  |
| **Hispanic Ethnicity** | 6 (2.4%) | 0 (0%) | 1 (1.1%) | 2 (4.2%) | 3 (6%) | 0.2 |
| **Insurance type** |  |  |  |  |  | <0.001 |
| Private | 106 (43%) | 13 (25%) | 15 (16%) | 30 (63%) | 48 (91%) |  |
| Public | 139 (57%) | 38 (75%) | 78 (84%) | 18 (38%) | 5 (9.4%) |  |
| **Marital status** |  |  |  |  |  | <0.001 |
| Married/Parter | 163 (67%) | 29 (57%) | 47 (51%) | 40 (83%) | 47 (89%) |  |
| Single | 82 (33%) | 22 (43%) | 46 (49%) | 8 (17%) | 6 (11%) |  |
| **Maternal Education** |  |  |  |  |  | <0.001 |
| ≤HS | 115 (47%) | 34 (67%) | 63 (68%) | 13 (27%) | 5 (9.4%) |  |
| >HS | 130 (53%) | 17 (33%) | 30 (32%) | 35 (73%) | 48 (91%) |  |
| **Family Income** |  |  |  |  |  | <0.001 |
| <\$25k | 88 (36%) | 27 (53%) | 49 (53%) | 10 (21%) | 2 (3.8%) | |
| \$25k-\$50k | 49 (20%) | 13 (25%) | 26 (28%) | 7 (15%) | 3 (5.7%) | |
| >\$50k | 108 (44%) | 11 (22%) | 18 (19%) | 31 (65%) | 48 (91%) | |
| **Infant sex** |  |  |  |  |  | 0.2 |
| Female | 127 (52%) | 31 (61%) | 50 (54%) | 24 (50%) | 22 (42%) |  |
| Male | 118 (48%) | 20 (39%) | 43 (46%) | 24 (50%) | 31 (58%) |  |
| **Delivery mode** |  |  |  |  |  | 0.14 |
| C Section | 94 (38%) | 16 (31%) | 30 (32%) | 23 (48%) | 25 (47%) |  |
| Vaginal | 151 (62%) | 35 (69%) | 63 (68%) | 25 (52%) | 28 (53%) |  |
| **Birthweight (kg)** | 3.29 (2.95, 3.53) | 3.27 (2.87, 3.46) | 3.22 (2.83, 3.50) | 3.32 (3.09, 3.62) | 3.37 (3.07, 3.61) | 0.2 |
| **Breastfeeding initiation** |  |  |  |  |  | 0.023 |
| Did not initiate BF | 33 (13%) | 10 (20%) | 18 (19%) | 3 (6.3%) | 2 (4%) |  |
| Initiated BF | 212 (87%) | 41 (80%) | 75 (81%) | 45 (94%) | 51 (96%) |  |
| **Days of BF** | 83 (16, 312) | 46 (9, 138) | 33 (2, 123) | 227 (47, 430) | 310 (122, 428) | <0.001 |

<sup>1</sup>Median (Q1, Q3); n (%)
<sup>2</sup>Kruskal-Wallis rank sum test; Pearson's Chi-squared test; Fisher's exact test

**Supplemental table 2:** Distribution of new serologically-identified RSV infections in 194 PREVAIL children.

|  |  | All new<br>concentration<br>change-identified<br>infections<br>*N*=133 | Method used to identify infection |  |  |
| --- | --- | --- | --- | --- | --- |
|  |  |  | IgA only<br>*n*=51 (38%) | IgG only<br>*n*=3 (2%) | Both<br>*n*=79 (60%) |
| **Age category** | < 6 months | 2 (2%) | 2 (4%) | 0 (0%) | 0 (0%) |
|  | 6-11 months | 11 (8%) | 9 (18%) | 0 (0%) | 2 (3%) |
|  | 12-17 months | 59 (44%) | 19 (37%) | 2 (67%) | 38 (48%) |
|  | 18-24 months | 61 (46%) | 21 (41%) | 1 (33%) | 39 (49%) |
| **Infection**<br>**number** | First | 57 (43%) | 21 (41%) | 0 (0%) | 36 (46%) |
|  | Second | 61 (46%) | 28 (55%) | 2 (66%) | 31 (39%) |
|  | Third | 15 (11%) | 2 (4%) | 1 (33%) | 12 (15%) |
| Legend:<br>Mixed effects classification and regression tree (CART) analysis was used to identify thresholds of positivity using pre-fusion F IgA and IgG assays from serum collected at 6 weeks and 6, 12, 18, and 24 months of age. CART-derived thresholds of >2.02 change in log<sub>10</sub> concentration of IgA or a >0.32 change in log<sub>10</sub> concentration of IgG were applied to PREVAIL participants who participated in the study at least 18 months and were either ≥70% adherent in weekly sample submission or provided a serum sample at 18 or 24 months of age. |  |  |  |  |  |

**Supplemental table 3:** Comparison between concentration change method and ROC-derived seropositivity in 194 evaluable children in the PREVAIL Cohort.

|  |  |  |  |  |
| --- | --- | --- | --- | --- |
|  |  | Concentration change<br>log<sub>10</sub> AU<br>IgA >0.202 or IgG>0.32 |  |  |
|  |  | Pos | Not Pos | % agreement |
| ROC-derived IgA seropositivity | Pos | 140 | 0 | 98.5% |
|  | Not pos | 3* | 51 |  |

Mixed effects classification and regression tree (CART) analysis was used to identify thresholds of positivity of >2.02 change in log<sub>10</sub> concentration of IgA or a >0.32 change in log<sub>10</sub> concentration of IgG as the most predictive threshold for use in identifying RSV infections. Seropositivity by the laboratory-determined ROC curves was limited to IgA, as 100% of children were seropositive by IgG from birth until age 6 months
- The three participants identified as seropositive using the CART-defined thresholds were positive by IgG only and confirmed as positive by IgG in the ROC analysis.

